# Does blood viscosity affect cerebral blood flow? A study in a population living in a high-altitude city

**DOI:** 10.1101/2023.01.15.23284577

**Authors:** Charles Huamaní, William Bayona-Pancorbo, William Sarmiento, Golda Córdova-Heredia, Luz Cruz-Huanca, Paulina Damián-Saavedra, Nathalie Requena, Víctor Oré-Montalvo, Carlos Pérez-Alviz, Juan C. Acuña-Mamani

**Affiliations:** Universidad Andina del Cusco. Cusco, Perú; Hospital Nacional Adolfo Guevara Velasco. Cusco, Perú; Universidad de San Martin de Porras. Lima, Perú

**Author notes:** **Corresponding author:** Charles Huamaní, MD, Mg. **Competing interests.** The authors declare no competing interests.

**Keywords:** viscosity, hemorheology, high-altitude, blood

## Abstract

**Background:** Viscosity affects flows by increasing resistance to movement; therefore, variations in blood viscosity (BV) levels could modify the cerebral blood flow.

**Objective:** This study aimed to determine the level of correlation between BV and cerebral blood flow in people acclimated to chronic hypoxia who have high BV levels.

**Methods:** Prospective observational study was conducted among clinically healthy young adults living in the city of Cusco (3,399 m above sea level). All participants were examined at low and high shear rates (75 and 300 sec^−1^) to simulate the dynamic component of BV. A transcranial Doppler study of the middle cerebral artery was performed to measure systolic, diastolic, and mean flow velocities (FVs) and resistance and pulsatility indices (PIs).

**Results:** A total of 131 participants were included. The median viscosity levels were 5.01cP [interquartile range (IQR): 4.45–5.73cP] at 300 sec^−1^ and 6.16 cP [IQR: 5.58-7.20 cP] at 75 sec^−1^, the mean FV was 57 m/s [IQR: 50–65 m/s^5^], and the PI was 0.91 [0.86–1.02]. BV was negatively correlated with mean FV (r: −0.17, p=0.007), while it showed no correlation with other values of blood flow, resistance, or PI.

**Conclusions:** Changes in BV levels have a minimal impact on the mean FV but not on other parameters. This finding suggests that in young and clinically healthy individuals, there are autoregulation mechanisms that compensate for BV variations, although they are not completely understood.

## Introduction

Blood viscosity (BV) indicates the resistance of blood when it is moving; thus, BV has been studied as a factor that could modify cerebral blood flow (CBF), although results have not been consistent in healthy people ^1^. This could be due to the methodologies used to measure CBF or BV, the populations selected, or the presence of autoregulatory mechanisms that allow CBF to remain stable despite variations in BV. By contrast, clinical studies indicated that cerebral small vessel infarction is correlated with elevated BV level, suggesting that it may have an influence on the BV ^2,3^. This finding suggests that it could have a negative influence on CBF, probably when autoregulatory mechanisms are decompensated.

These inconsistent results could also be due to the physics and dynamics of blood as a fluid; the blood is composed of several heterogeneous components, such as red blood cells, platelets, proteins, etc., which means that its behaviour as a fluid does not obey Newtonian laws. ^4,5^. At different tensions, the BV will change; therefore, during systole or diastole or when passing through a large vessel or capillary, the BV level varies, which makes it different from other haematological parameters that are usually stable. This aspect, which has been widely studied from the rheology and physics point of view, has not been completely ^6-8^ and fully integrated into the field of clinical study.

In order to provide a middle ground between a healthy and a diseased settler, our study aimed to assess settlers whose physiological self-regulatory mechanisms have already been exhausted. Hence, young and clinically healthy people were included in this study, who were expected to show preservation in their self-regulatory mechanisms, but who live in a city at high altitude and are exposed to physiological challenges, acclimatisation, and chronic hypoxia ^9^. The main compensatory mechanism consists of increasing the haemoglobin levels, which increases the BV values above those observed in people living at sea level ^10^. This study aimed to evaluate the variations in BV levels and to determine their correlation with some parameters of CBF.

## Methods

### Design and population

An observational, analytical and prospective study was carried out in the city of Cusco, located 3,399 m above sea level ^11^. Clinically healthy adults aged 18–40 years were invited to participate in the study. Approximately 70% of the participants indicated that they had lived in Cusco all their lives, while the rest had lived in Cusco for more than 6 months. This was a criterion for inclusion in the study because it was an appropriate period to assess for any physiological changes that may occur due to exposure to a high altitude ^9^. People who consumed non-steroidal anti-inflammatory drugs, anticoagulants, alcohol, or tobacco during the last 72 hours prior to the evaluations were excluded.

The study of Grotta, Ackerman, Correia, Fallick and Chang ^12^ evaluated the correlation of BV and CBF; they used a sample size of 53 patients, obtaining a correlation of −0.35 (p=0.01). Based on these data and with a significance level of 0.01 and a statistical power of 90%, a sample size of 118 participants would be required. Given that the temporal window (access to perform an adequate transcranial Doppler approach) is inadequate in 10% of the population, a total of 130 participants were included in the study ^13^. However, as each person provides two windows of access for assessment, this would increase to no less than 420 records; hence, the statistical power would be no less than 90% for a significance level of 0.001.

### Blood viscosity measurement

After obtaining informed consent, 3 ml of venous blood sample was collected from all eligible patients while seated on a chair with the left arm flexed and was mixed with ethylenediaminetetraacetic acid. This procedure was performed following the respective biosafety guidelines for handling specimens to avoid the spread of coronavirus disease 2019 (COVID-19).

Approximately 0.5 ml of venous blood sample was used, which was processed at a controlled temperature of 37°C, in the Brookfield AMETEK brand viscometer, model DV2T LV CP (cone-plate), and BV values were expressed in poises (P); the measurement was performed in accordance with the existing regulations ^14,15^.

Viscosity measurements in fluids, such as blood, need to be assessed in different scenarios due to its behaviour as a non-Newtonian fluid ^4,5^. Therefore, the study of Kim, Cho, Lee, Park, Moon, Hur, Kim and Yun ^16^ proposed the use of differential measurements to calculate the BV level using 1 and 300 sec^−1^ as *share rate* through a scanning capillary tube viscometer. For the process to be replicable in a rotational viscometer, the revolutions per minute (RPMs) and its normalisation were adjusted according to the *share rate* of the spindle used. Therefore, after performing the standardisation tests on whole blood samples, the measurements were performed at 10, 20, 30, and 40 RPM, which are equivalent to 75, 150, 225, and 300 sec^−1^. The extremes represent the highest and lowest VS values, respectively, while the intermediate values determines the reliability of the measurements by assessing a decreasing variation.

### Transcranial Doppler study

The transcranial Doppler ultrasound was performed on the same day as the viscosity sampling, following the established procedures for assessing and identifying the middle cerebral artery, with a time difference of up to 3 hours. Transcranial Doppler ultrasound is noninvasive, provides rapid results, and obtains FVs that are consistent with the invasively or directly measured flow measurements ^17^.

Transcranial Doppler ultrasound with insonation was performed using a 2-MHz probe inserted through the temporal window; the probe was advanced at 50-mm depth until the arterial flow whose direction is close to the probe was identified ^18^. Each middle cerebral artery (left and right) was insonated, and a velocity difference in systolic flows of not greater than 10 cm/s was considered valid. In each artery, the systolic, diastolic, and mean flow velocities (FVs), as well as the resistance index and pulsatility index (PI) were obtained ^19^. Since the transcranial Doppler ultrasound is a dynamic study, which presents fluctuations due to various conditions or manoeuvres, the study was performed in stable patients and at rest for no less than 10 minutes.

### Data processing

The data obtained were entered and exported to the statistical programme STATA/IC 16, and the variables were coded, revised, and analysed. The statistical analysis included the description of the study variables, using absolute and relative frequencies, as well as medians with interquartile ranges [IQRs].

The correlation between BV values and CBF parameters was evaluated using the Spearman’s correlation coefficients (R-value) because these parameters do not follow a normal distribution. A p value of <0.05 were considered significant.

### Ethical

The study was conducted in a clinically healthy adult population, and informed consent was obtained from all participants; the study was approved by the ethics and research committee of the Hospital Nacional Adolfo Guevara Velasco, EsSalud Cusco (resolution no. 419-GRACU-ESSALUD-2020). Due to the COVID-19 pandemic contingency, all participants were required to wear masks and face shields and complied with social distancing and other standard biosecurity measures, minimising exposure time during scheduled appointments.

## Results

A total of 131 volunteers were included, 78 (59.5%) were women, with a median age of 24 [IQR: 22–29] years. The median haemoglobin level was 16.1 mg/dl [IQR: 14.8–17.9], with variations between men (18.1 mg/dl, IQR: 17.5–18.7) and women (15.3 mg/dl, IQR: 14.1–16.0). The other haematological parameters measured are presented in Table 1.

**Table 1.** Characteristics of healthy high-altitude residents included in the study

|  | Total<br>N° 131 (100%) | Males<br>N° 53 (40.5%) | Women<br>N° 78 (59.5%) |
| --- | --- | --- | --- |
| Age (years) | 24 [22-29] | 25 [23-30] | 24 [21-28] |
| Hemoglobin (g/dl) | 16.1 [14.8-17.9] | 18.1 [17.5-18.7] | 15.3 [14.1-16.0] |
| Hematocrit (%) | 47.9 [44.0-52.8] | 53.1 [51.3-54.9] | 45.2 [42.4-47.8] |
| Triglycerides (mg/dl) | 100 [78 - 167] | 131 [87 - 209] | 96 [67 - 137] |
| Total cholesterol (mg/dl) | 185 [161 - 212] | 192 [170 - 214] | 180 [157 - 208] |
| HDL (mg/dl) | 53 [47 - 58] | 53 [46 - 58] | 53 [48 - 59] |
| LDL (mg/dl) | 105 [86 - 136] | 112 [88 - 142] | 99 [84 - 131] |
| Total proteins (g/dl) | 7.69 [7.36-8.06] | 7.91 [7.61-8.18] | 7.56 [7.27-7.87] |
| Albumin (g/dl) | 4.59 [4.39-4.80] | 4.75 [4.59-4.87] | 4.50 [4.33-4.69] |
| Globulin (g/dl) | 3.10 [2.89-3.37] | 3.17 [2.93-3.45] | 3.07 [2.87-3.28] |
| Red blood cells (10<sup>6</sup>/uL) | 5.29 [4.93-5.74] | 5.77 [5.58-6.05] | 5.01 [4.74-5.29] |
| White blood cells (10<sup>3</sup>/uL) | 6.11 [5.15-6.89] | 5.90 [5.18-6.63] | 6.42 [5.14-7.14] |
| Platelets (10<sup>3</sup>/uL) | 291 [259 - 330] | 274 [246 - 301] | 302 [259 - 355] |
| Glucose (mg/dl) | 89 [85 - 95] | 93 [86 - 100] | 87 [84 - 94] |

HDL: high-density lipoprotein, LDL: low-density lipoprotein.
Values between [ ] correspond to the interquartile range.

The blood viscosity levels ranged from 5.01 cP [IQR 4.45–5.73cP] at 300 sec^−1^ to 6.16 cP [5.58-7.20 cP] at 75 sec^−1^ (Table 2). Meanwhile, the mean CBF velocity was 57 cm/s [IQR: 50–65], and the PI was 0.9 [0.86–1.02].

**Table 2.** Blood viscosity and cerebral blood flow values in healthy high-altitude residents

|  | Total<br>N° 131 (100%) | Males<br>N° 53 (40.5%) | Women<br>N° 78 (59.5%) |
| --- | --- | --- | --- |
| Blood viscosity (cP) |  |  |  |
| 75 sec<sup>-1</sup> | 6.16 [5.58-7.20] | 7.24 [6.87-7.89] | 5.70 [5.27-6.10] |
| 150 sec<sup>-1</sup> | 5.41 [4.89-6.32] | 6.33 [5.89-6.87] | 4.97 [4.69-5.41] |
| 225 sec<sup>-1</sup> | 5.14 [4.60-5.92] | 6.09 [5.57-6.46] | 4.72 [4.40-5.12] |
| 300 sec<sup>-1</sup> | 5.01 [4.45-5.73] | 6.02 [5.44-6.33] | 4.58 [4.32-4.99] |
| Cerebral blood flow |  |  |  |
| Systolic velocity (cm/s) | 89 [80-100] | 87 [74-98] | 89 [81-101] |
| Diastolic velocity (cm/s) | 37 [31-42] | 35 [29-41] | 38 [32-43] |
| Mean velocity (cm/s) | 57 [50-65] | 55 [47-62] | 59 [53-65] |
| Resistance index | 0.85 [0.55-0.63] | 0.59 [0.55-0.65] | 0.58 [0.55-0.62] |
| Pulsatility index | 0.91 [0.86-1.02] | 0.94 [0.86-1.08] | 0.89 [0.82-0.98] |

Values between [ ] correspond to the interquartile range.

The BV levels had a weak and negative correlation with mean CBF velocity, but not with the other parameters evaluated, with small variations according to the viscosity measurement, ranging from a value of r = −0.132 at 75 sec^−1^ to a value of r = −0.170 at 300 sec^−1^ (Table 3).

**Table 3.** Correlation between blood viscosity and CBF in healthy high-altitude residents

|  | Blood viscosity |  |  |  |
| --- | --- | --- | --- | --- |
|  | 75 sec<sup>-1</sup> | 150 sec<sup>-1</sup> | 225 sec<sup>-1</sup> | 300 sec<sup>-1</sup> |
| Mean velocity | r: -0.132<br>**p: 0.037** | r: -0.124<br>**p: 0.049** | r: -0.147<br>**p: 0.019** | r: -0.170<br>**p: 0.007** |
| Systolic velocity | r: -0.092<br>p: 0.148 | r: -0.094<br>p: 0.136 | r: -0.119<br>p: 0.059 | r: -0.136<br>**p: 0.031** |
| Diastolic velocity | r: -0.065<br>p: 0.302 | r: -0.048<br>p: 0.449 | r: -0.063<br>p: 0.322 | r: -0.079<br>p: 0.212 |
| Pulsatility index | r: 0.065<br>p: 0.304 | r: 0.032<br>p: 0.613 | r: 0.031<br>p: 0.626 | r: 0.035<br>p: 0.581 |
| Resistance index | r: 0.028<br>p: 0.662 | r: -0.002<br>p: 0.981 | r: -0.004<br>p: 0.950 | r: -0.003<br>p: 0.963 |

## Discussion

Our study has some strengths. It included an understudied population, expanded the sample size of previous studies, and used several levels of BV following the current recommendations for its measurement ^15^. Initially, our results may not seem to provide a contribution, due to the low correlation identified in only one of the parameters evaluated. However, the information must be evaluated in the context in which it is presented. First, the study was conducted in a young population and in those with no symptoms or without described diseases; hence, their autoregulatory mechanisms should be unscathed and should correctly handle hemodynamic alterations to which they have been exposed to. Second, they have been chronically exposed to hypoxia, as evidenced by their elevated haemoglobin levels, while they must have undergone other physiological changes to acclimatise. Third, the sample size was extremely large. According to the estimates, the statistical power was superior to that reported in previous studies. Fourth, despite the previous points, a small impact on CBF was identified.

The association found, although small and only for mean FV, suggests that BV has an impact on CBF, but it is compensated by the self-regulatory mechanisms present as this is a young population ^20^. In this regard, we have to analyse that the systolic and diastolic FVs are pulsatile components of CBF, while the mean flow velocity is a continuous component and calculated based on the systolic and diastolic flow velocity values ^21^. These aspects should be interpreted with caution because the absence of association between BV and systolic and diastolic values, but the presence of association between BV and mean flow values, suggests a regulation for continuous values, but not for pulsatile values; this may be because the variations of BV at this level are relatively small and because the mechanisms of autoreactivity or vasoreactivity also remain constant when cerebral perfusion is adequate ^22,23^.

Studies in individuals who ascend rapidly to high altitudes show that CBF is altered by acute exposure to high altitude; in individuals who waited until acclimatisation to high-altitude areas resulted in hypoxia, a dynamic modification process of CBF was observed ^24,25^. In individuals who waited until acclimatisation to high-altitude areas resulted in hypoxia, dynamic modification of CBF due to autoregulation was observed; however, this mechanism is not completely understood, and it may not have an impact on the CBF ^26^ and may not have a significant impact on brain physiology ^27^. However, in all studies analysed, the effect of BV, which also varies according to acclimatisation status, was not evaluated. Our study, however, suggests that BV has a small impact on CBF dynamics and it is related to the BV reported in chronic high-altitude dwellers. Although our results are limited, if we incorporate them to those obtained in clinical settings, we could suggest that BV is related to BV in chronic high-altitude residents ^2,3^. Although our results are limited, if we incorporate them to those obtained in clinical settings, we could suggest that BV does play a role in the alterations of CBF levels and could be manifested when cerebral autoregulation mechanisms are overcome, although these have a lower expression in young and healthy people.

Although the effect of BV on CBF has not been studied further, other investigations have evaluated the impact of haematocrit on cerebral autoregulation, finding an association between higher haematocrit levels and alterations in CBF levels ^28,29^ and have even modelled these phenomena ^30,31^. Our population has higher BV and haematocrit levels than those living at sea level, but without demonstrating excessive erythrocytosis (men: Hb ≥20 mg/dl, women: Hb ≥18 mg/dl) ^32^. Moreover, our results are consistent with those of other evaluations conducted in populations living in high-altitude areas ^10^. The drawback of these indirect approaches is that BV is a dynamic phenomenon; therefore, even if we maintain a constant haematocrit, the BV level will change; hence, the impact of BV is probably not significant.

The low impact identified in our study could be attributed to the method we used, as previous studies developed several methods for measuring the CBF and some processes for measuring BV were not standardised ^23^. Transcranial Doppler ultrasound was performed, which is a cerebral perfusion study and measures some components of blood flow ^1,33^. It measures some components of CBF and has a good correlation with direct or invasive studies ^17^. The association between BV and CBF has been investigated since the studies of Grotta, Ackerman, Correia, Fallick and Chang ^12^ and Brown and Marshall ^34^. Although they reported that these two factors had a positive correlation, they presented technical limitations for both BV and CBF measurements due to the technological development that occurred during that time. Grotta, Ackerman, Correia, Fallick and Chang ^12^ did not directly measure the BV levels, but estimated its value using a formula based on the haematocrit and fibrinogen levels; meanwhile, Brown and Marshall ^34^ measured CBF using xenon-133. These variations, in addition to the use of small sample size, may explain why other studies have shown inconsistent data ^1^.

In addition, BV levels are variable according to the share rate to which they are subjected, but this is not the only factor that could alter the BV levels. A component that is usually mentioned is the calibre of the vessel through which the blood transits, although BV can vary significantly according to this calibre; this is limited to vessels with less than 0.3 mm in diameter (arterioles and capillaries), which is known as the Fahræus-Lindqvist effect ^35^. In our case, we evaluated the middle cerebral artery, whose average diameter is theoretically 3.2 mm, although the diameter can vary in patients with hypocapnia or hypercapnia ^36^. These changes would have no impact on changes in BV levels. Although our results are only limited only to the large arteries and do not provide details on the events that can occur at the arteriole or capillary level, the physiological impact of BV on small vessels remain controversial ^35^. Clinically, this impact is associated with small vessel infarctions ^2^.

### Limitations

Our study focused mainly on the basic science and physiology; hence, only young patients were included, but this limited selection might not explain all the changes that happened in older age settlers. Hence, the extension of the study is required, and the progressive loss of autoregulatory mechanisms should be assessed. Additionally, the transcranial Doppler ultrasound was used for the measurement of CBF, even though several additional techniques can be used to determine other parameters of greater utility, such as cerebral blood perfusion or autoregulation; however, there is no standardised method for measuring CBF ^37^. Moreover, there is no standard method for reporting CBF in research. Finally, our study only allows the assessment of medium to large calibre arteries; in small calibre arteries, the physics of viscosity has a different behaviour, and these variations could be the cause of the variations in CBF ^35^. Moreover, these variations could be the cause of the negative clinical impact.

## Conclusion

Only a slight variation was observed in BV levels. However, this variation had a significant impact on mean flows, but not on the other parameters. This indicates that, in young and clinically healthy individuals, there are autoregulatory mechanisms that compensate for BV variations, although it is not completely understood. This could play a role in the pathogenesis of cerebrovascular diseases that requires further clinical studies in older population.

## Data Availability

All data produced in the present study are available upon reasonable request to the authors.

